# CXCL14 suppresses the progression of colon cancer by regulating tumor epithelial-mesenchymal transition and tumor microenvironment

**DOI:** 10.1101/2024.02.26.24303307

**Authors:** Yinjie Zhang, Yue Jin, Siyi Wang, Yuchen Niu, Buyong Ma, Jingjing Li

## Abstract

**Background:** The widespread silencing of CXCL14 in advanced colon cancer underscores the association between CXCL14 and the development of colon cancer. Some studies have demonstrated *in vitro* that CXCL14 can inhibit the growth and metastasis of colon cancer cell, and it has also been studied in other disease models for its regulation of immune cell infiltration.

**Aims:** The aim of this study is to clarify the transcriptional regulation of colon cancer cells mediated by CXCL14 and its regulatory role in the tumor microenvironment.

**Method:** We analyzed the expression characteristics of CXCL14 in clinical databases of colon cancer. CXCL14 overexpression cell lines were established to study its functions on gene transcriptional regulation and cell physiology. Through subcutaneous tumor models, we investigated the effects of CXCL14 on the immune microenvironment.

**Result:** Firstly, the clinical data revealed that CXCL14 is silenced during cancer progression, and negatively associated with EMT and cell proliferation markers, and positively associated with the abundance of T cells and NK cells in colon tissue. Secondly, RNA-seq reveals that CXCL14 overexpression is negatively associated with cell EMT, and inhibits tumor migration and invasion. Western blot (WB) assay confirmed that CXCL14 inhibits the Erk/MAPK and Akt signaling pathways. Finally, subcutaneous tumor models demonstrate that CXCL14 overexpression inhibited tumor growth, increased the infiltration of tumor-associated T cells and DC cells, activated the anti-tumor immune response, antigen processing and presentation, and T helper differentiation in the tumor microenvironment.

**Conclusion:** CXCL14 becomes silenced in late-stage colon cancer samples. *In vivo* and *in vitro* experiments demonstrate that CXCL14 inhibits tumor EMT, proliferation, and metastasis through autocrine signaling. It also enhances the infiltration of T and NK lymphocytes through paracrine signaling and inhibits the proliferation of subcutaneous tumors.

## Background

CXCL14 (also known as breast and kidney expressed chemokine, BRAK) is a non-ELR CXC chemokine. It is highly conserved in mammals, with only two amino acid differences between mice and humans. CXCL14 is constitutively expressed in several normal tissues, including adipose tissue, brain, breast, uterine cervix, lung, kidney, and skin. CXCL14 has multiple biological functions, including involvement in the regulation of cancer, immune response, and epithelial cell proliferation and migration. It is an effective inhibitor of angiogenesis[1] and an antimicrobial agent [2]. Previous studies have shown that CXCL14 exhibits chemotactic activity for monocytes [3]. Additionally, CXCL14 specifically increases the chemotaxis of CD56+ natural killer (NK) cells [4]. CXCL14 is involved in the regulation of many biological processes *in vivo*, including inflammatory immune response, tumor-associated angiogenesis, activation of tumor-specific immunity by the host, and autocrine regulation of tumor growth.

The widespread silencing of CXCL14 in clinical colon cancer tissues suggests its potential tumor suppressor function [5], This tumor suppressor function may be derived from the multifaceted functions of CXCL14.

Firstly, CXCL14 has the potential to inhibit the motility and migration of tumor cells. Research has shown that it can inhibit the migratory activity of human colon cancer cells *in vitro*[6, 7]. Furthermore, CXCL14 also affects the motility of other cells, including trophoblast outgrowth through autocrine and paracrine forms[8], and it regulates neural cell migration [9]. Therefore, CXCL14 may be associated with epithelial-mesenchymal transition (EMT). Secondly, CXCL14 may play a role in regulating the tumor angiogenesis microenvironment. Research has shown that CXCL14 can inhibit angiogenesis in vitro in human liver cancer cells [1]. Additionally, CXCL14 can also inhibit angiogenesis in corneal and dental pulp tissues in mice [10]. Finally, CXCL14 plays a complex role in the immune microenvironment of colon cancer. CXCL14 may possess anti-tumor effects. CXCL14 can attract and activate NK cells and macrophages [3, 4], enriching them in the tumor microenvironment, thereby enhancing the regulation of anti-tumor immune responses against these cells. The silencing of CXCL14 during the growth of colon cancer suggests that the unaltered expression of CXCL14 may pose an obstacle to the development of this cancer[5]. Additionally, CXCL14 may be involved in immunosuppression. Within the tumor’s microenvironment, CXCL14 can indirectly suppress immune responses by modifying the function and/or recruitment of Treg cells[11]. This interference may aid tumor cells in evading attacks from the immune system.

The molecular mechanism of CXCL14 remains enigmatic, with its receptor still unidentified. While some studies propose that CXCR4 may serve as the natural receptor for CXCL14, contradictory research findings [12–15] have impeded progress in comprehending CXCL14. Nevertheless, recent studies have revealed that CXCL14 can engage the MRGPRX2 receptor[16], offering valuable insights into its molecular mechanism. Additionally, CXCL14 exhibits synergistic effects with various ligands of the same family [17], possibly through the formation of heterodimers [18].

In summary, more research is required to fully comprehend the role of CXCL14 in the immune microenvironment of colon cancer. The function of this chemokine may extend beyond the direct and indirect immune regulation described above, encompassing additional facets that remain unexplored. Studying the function and mechanism of CXCL14 will not only enhance our understanding of the etiology and progression of colon cancer but also offer novel insights for the development of immunotherapies for solid tumors. The recruitment of immune cells by CXCL14 may offer new therapeutic targets for combating tumor growth.

Through the establishment of a mouse CXCL14 overexpression cell line, this study aimed to investigate the impact of CXCL14 on the proliferation, migration, invasion, and other cellular behaviors of colon cancer cells. Additionally, it aimed to unravel the regulatory mechanism of CXCL14 on cell signaling pathways using transcriptomics. In addition, the study has also revealed the regulatory mechanism of CXCL14 on cell signaling pathways through transcriptomics. Furthermore, the interaction between CXCL14 and the immune system in vivo has been clarified through subcutaneous tumor xenografts and tumor transcriptomics.

## Methods and materials

### 1. Cell culture

MC38 cells (ATCC CRL-2639) were cultured in DMEM medium supplemented with 10% FBS, while CT26 (ATCC CRL-2638) and HCT15 cells (CCL-225) were cultured in RPMI1640 medium with 10% FBS. All cells were maintained in a humidified incubator at 37°C with 5% CO2 and 80% humidity.

### 2. Stable cell line construction

To generate a MC38 cell line inducibly expressing CXCL14, we constructed a doxycycline-inducible lentiviral vector for CXCL14. The open reading frame of mouse CXCL14 and an internal ribosome entry site (IRES) were cloned into the pLVX-TetOne-EGFP vector (Addgene, plasmid #171123), generating the pLVX-TetOne-CXCL14 construct (plasmid sequence in Supplementary Material 1). The lentiviral particles were generated by transfecting the TetOne-CXCL14 plasmid, psPAX2 (Addgene #12260), and pMD2.G (Addgene #12259) into 293T cells (ATCC and CRL-3216) using Lipofectamine 2000 reagent (Invitrogen, 11668027). The virus particles were then concentrated using Lenti-X Concentrator (TAKARA, 631231). The virus particles were used to infect MC38 cells, and 48 hours later, puromycin (2 ug/ml) was added to select for infected cells. Single clones were then isolated and induced with 1 ug/mL doxycycline to express CXCL14. The clones with green fluorescence were selected and further characterized by quantitative PCR to confirm overexpression of CXCL14 (primers are listed in Table S1).

CT26 stable cell line construction: The open reading frame of mouse CXCL14 gene was cloned into the pcDNA3.1 vector (plasmid sequence in Supplementary Material 2). Single clones were selected using 200 ug/mL G418 and confirmed for overexpression of CXCL14 using quantitative PCR (primers are listed in Table S1).

HCT15 stable cell line construction: The pHIV-zsgreen plasmid (Addgene #18121) was modified by replacing the zsgreen gene with a puromycin resistance gene (puromycin N-acetyltransferase). The human CXCL14 gene open reading frame was then cloned into the multi-cloning site of the modified pHIV-zsgreen plasmid (plasmid map in Supplementary Material 3). The plasmid was transfected into HCT15 cells, and single clones were selected using 2 ug/ml puromycin. Overexpression of CXCL14 was confirmed using quantitative PCR (primers are listed in Table S1).

### 3. Subcutaneous tumorigenesis in mice

7-week-old male mice were used for tumorigenesis. The tumor cells were cultured in complete medium until confluent (80-90%), digested with trypsin-EDTA, washed twice with HBSS, resuspended in PBS and adjusted to a density of 10^7^/mL. Using a 1 mL syringe, the tumor cells were injected subcutaneously at the right axilla, with 100 uL of cell suspension (containing 10^6^ cells) injected into each mouse. For the tumor proliferation experiment, mice were fed with sterile water containing 1 mg/mL DOX after subcutaneous tumorigenesis, and tumor size and mouse weight were measured every two days until the end of the experiment. For the study of CXCL14 regulation of the tumor microenvironment, mice were given free access to 1 mg/mL DOX when the tumor grew to about 300 mm^3^, and tumor tissue was collected for further detection after 3 days.

### 4. RNA-seq

Total RNA was extracted from 10 cm dishes of cells or 0.1 g of tumor tissue using column purification (TAKARA, 9767). RNA-seq experiments were performed by Beijing Geneplus Technology Co. Ltd. CXCL14 overexpression tumor cell samples included control and CXCL14 overexpression groups, with two biological replicates per group. Tumor samples included control and CXCL14 induction groups, with four biological replicates per group. Each sample was sequenced at a depth of 10G.

### 5. Quantitative PCR

Total RNA was extracted using Trizol (Invitrogen, 15596026) combined with centrifuge column method (Sangon, B615025). mRNA was reverse transcribed to cDNA (Novozymes, R302-01). Using Sybrgreen quantitative PCR reagents (TAKARA, RR820Q), gene expression was measured by ΔΔCt method. Quantitative PCR primers are shown in Table S1).

### 6. Western blot

The tissue and cells are lysed in a lysis buffer containing protease inhibitors and phosphatase inhibitors. The protein concentration is determined using a BCA assay kit. The proteins are denatured in an SDS sample buffer by boiling. Equal amounts of proteins are separated by SDS-PAGE, and then transferred onto a nitrocellulose membrane (Millipore). The membrane is incubated with a primary antibody overnight at 4°C. The house-keeping gene GAPDH is detected by an anti-beta-actin antibody as an internal control. The membrane is then incubated with the corresponding secondary antibody conjugated to horseradish peroxidase at room temperature for 1 hour, and the proteins are visualized using a chemiluminescent substrate. The gel is scanned using a Biorad gel imaging system, and the bands are quantitated using Image J software.

### 7. Invasion experiment

50 μL of 2 mg/ml Matrigel is used to coat the transwell invasion chamber, and the edges of the cell plate are sealed with sealing film. The invasion chamber is placed at 4°C overnight. The invasion chamber is washed twice with cold PBS, and 10^5 cells in 100 μL of serum-free medium are added to the upper compartment of the transwell, while 600 μL of complete medium containing 10% FBS is added to the lower compartment. The cells are cultured at 37°C for 24 hours. The cells in the lower compartment of the invasion chamber are fixed with 4% paraformaldehyde for 10 minutes, stained with 1% crystal violet stain, and then photographed. Five random fields of view at 10x magnification are selected, and the number of cells that have invaded the lower compartment of the invasion chamber is counted.

### 8. Wound repair assay

Cells are seeded at a density of 100% confluence in a 12-well plate and cultured overnight until a monolayer of cells is formed. Multiple scratch lines are made on the monolayer cells using a 10uL pipette tip, perpendicular to the plate. The cells are then washed twice with PBS and cultured in medium containing 2% FBS. Microscopic images are taken at 0, 24, and 48 hours. The ImageJ software (version 1.53T) is used to analyze the cell migration distance. T-tests are used to analyze the differences in migration speed.

### 9. Histopathology and immunohistochemistry staining

Mice were sacrificed by cervical dislocation, and the tumor tissues were isolated after 70% ethanol disinfection. The tissues were fixed in 4% paraformaldehyde solution for 24 hours, embedded in paraffin, and sectioned into 5μm slices. The slices were stained with hematoxylin and eosin (HE) using standard protocol. The paraffin sections were stained with H-E using a staining kit (Yeasen, 60524ES60). The immunohistochemistry staining was performed using standard methods, including dewaxing, rehydration, antigen retrieval by heat treatment with 1% citric acid, and staining with antibodies against CD3 (ebioscience, 14-0030-82) and CD86 (ebioscience, 14-0862-82).

### 10. Flow cytometry detection

Tumor tissues are chopped into small tissue pieces (approximately 1mm3) using ophthalmic scissors. Five times the volume of the tumor tissue is added to the digestion solution (1% collagenase II + 100U/mL Hyalurondase + 0.1% DNAse + DMEM/F12), and the mixture is incubated at 37°C for 1 hour, with gentle mixing every 15 minutes. After digestion, the cells are filtered through a cell strainer to obtain a single-cell suspension. The cells are washed twice with five times the initial volume of PBS. The cell suspension is blocked with PBS containing 1% FBS for 10 minutes; Cells are adjusted to a concentration of 1×10^7^/ml in suspension, and 0.1 mL is transferred to an EP tube. Fc receptors are blocked by incubating with 0.5-1 μg of purified monoclonal antibody against mouse CD16/32 (E-AB-F0997A) at room temperature for 10 minutes. T lymphocyte subpopulations are stained using antibodies against CD3 (pc5.5, eBioscience, 45-0031-82), CD4 (FITC, eBioscience, 11-0041-82), and CD8 (APC, eBioscience, 17-0081-82). For macrophage staining, double staining is performed using F4/80 (APC, eBioscience, 17-4801-82) and CD11b (PC7, eBioscience, 25-0112-82). The recommended amount of fluorescently labeled antibodies is added according to the manufacturer’s instructions, and the antibodies are diluted in flow cytometry staining buffer (PBS containing 1% BSA). The mixture is vortexed and incubated in the dark at 4°C for 30 minutes. After washing the cells twice with 1 mL of staining buffer for 5 minutes each, centrifuging the cell suspension at 300g for 5 minutes, and discarding the supernatant, 0.2 mL of staining buffer is added to resuspend the cells. The cells are then analyzed and detected using a flow cytometer. Fixation with 1% paraformaldehyde is performed for 10 minutes, and the cells are then analyzed using the flow cytometer.

### 11. Data mining and analysis

The cBioportal database is utilized for the analysis and retrieval of expression profile data regarding colon cancer. The “Co-expression” feature aids in the retrieval of the correlation between the expression levels of other genes and the CXCL14 gene in datasets TCGA (PanCancer Atlas), Sidra-LUMC AC-ICAM (Nat Med 2023), CPTAC-2 Prospective (Cell 2019) and MSK (Nature Medicine 2022). The Gepia database was used to search for gene sets closely associated with CXCL14 expression in the normal colon tissue expression profile database Genotype-Tissue Expression (GTex). The “Similar Genes Detection” function provided by Gepia was used for retrieval. The GO and KEGG enrichment analysis of RNA-seq data was carried out using the online analysis system provided by JYJCloud (https://www.jyjcloud.com/), and the self-built gene set GO enrichment analysis was conducted on the official website of The Gene Ontology Resource[19]. Correlation analysis and t-test were performed using IBM SPSS software (Version 25). The Pearson correlation coefficient was used to represent the correlation of gene expression levels. T-test was performed with two-tail model.

## Result

### 1. Silencing of the CXCL14 gene is associated with poor prognosis in clinical cases

We conducted an analysis on three major colon cancer datasets (TCGA, CPTAC, and Sidra-LUMC) using the cbioportal database. The results revealed a negative correlation between CXCL14 expression and the stage of colon cancer (figure 1A), consistent with previous studies on the epigenetic silencing of CXCL14 [5]. The pathological sections of clinical samples indicated that CXCL14 is silenced in tissues of colon cancer and rectal cancer (figure 1B). Furthermore, common human and mouse colorectal cancer cell lines also exhibit almost complete silencing of CXCL14 (figure 1C, D). Similarly, in the three aforementioned databases, CXCL14 exhibits a significant negative correlation with several crucial EMT-related genes [20]. This finding suggests that CXCL14 may play a role in inhibiting tumor cell EMT phenotypes (p < 0.01, figure 1E, Supplementary Data 1). Including the three databases mentioned above, as well as the MSK dataset, all demonstrate a negative correlation between CXCL14 expression and the tumor proliferation marker TK1 in colon cancer cells (p < 0.001), indicating that CXCL14 is inversely associated with tumor proliferation (figure 1F, Supplementary Data 2).

**Figure 1.**
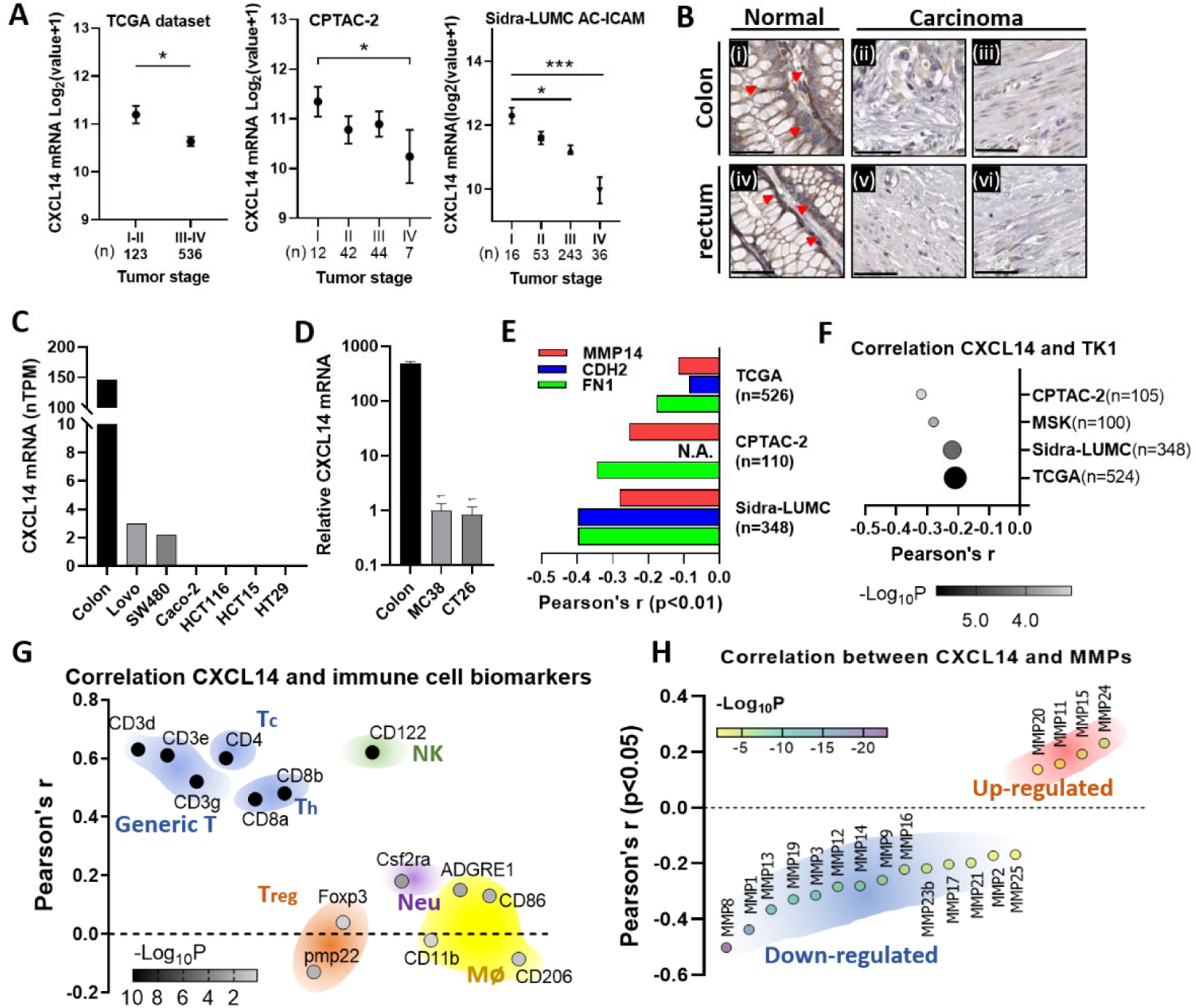
Gene silencing of CXCL14 is associated with adverse prognosis. (**A**) The colon cancer database showed that the expression level of CXCL14 is progressively downregulated during the development and metastasis of colon cancer. The data are represented by mean + SEM. **(B)** The ProteinAtlas database demonstrates that CXCL14 is highly expressed in normal colon (i, PID3753) and rectal gland cells (arrow) (iv, PID:3243), but its expression is significantly reduced in colon adenocarcinoma tissue (ii) PID:2002 and (iii) PID:1958 and rectal adenocarcinoma (v) PID:1424 and (vi) PID:1416 tissue. The bar represents 50 μM. **(C)** A comparison of CXCL14 expression levels in normal colon tissue and colon cancer cell lines. Normal colon tissue data were obtained from the GTEx database, while tumor tissue data were obtained from the Ensembl database. It was observed that several commonly used colon cancer cell lines exhibit basal silencing of CXCL14. **(D)** Quantitative PCR results for CXCL14 in mouse colon tissue and cell lines. The results indicate that the expression levels of MC38 and CT26 cells are extremely low. **(E)** The clinical database reveals a significant negative correlation between CXCL14 and EMT markers. CXCL14 exhibits significant negative correlations with three markers across three colon cancer databases. N.A. indicates that there is no data available for that marker in the database, while the remaining data exhibit a significant correlation coefficient (p<0.01). **(F)** CXCL14 exhibits a general negative correlation with the tumor cell proliferation marker TK1 in the four colorectal carcinoma database. **(G)** The correlation between CXCL14 and immune cell markers. According to the GTEx colon tissue expression profile database, CXCL14 exhibits a strong correlation with T cells markers (0.4<r<0.7), and a strong correlation with the NK cell marker CD122 (r>0.6). However, there is either no significant correlation or a weak correlation between CXCL14 and Treg cells, neutrophils, and macrophage markers (r<0.2). **(H)** Analysis of the correlation between CXCL14 expression levels and the expression levels of MMPs family genes in the Sidra-LUMC database.

We conducted an analysis on three major colon cancer datasets (TCGA, CPTAC, and Sidra-LUMC) on the cbioportal database. The results showed that CXCL14 expression is negatively associated with the stage of colon cancer (figure 1A), consistent with previous studies on the epigenetic silencing of CXCL14 [5]. The pathological sections of clinical samples showed that CXCL14 is silenced in tissues of colon cancer and rectal cancer (figure 1B). In common human and mouse colorectal cancer cell lines, CXCL14 is also almost silent (figure 1C, D). Similarly, in the three databases mentioned above, CXCL14 shows a significant negative correlation with several important EMT-related genes [20]. This result suggests that CXCL14 may be involved in the process of inhibiting tumor cell EMT phenotypes (p < 0.01, figure 1E, Supple data 1). Including the three databases mentioned above, as well as the MSK dataset, all show a negative correlation between CXCL14 expression and the tumor proliferation marker TK1 in colon cancer cells (p < 0.001), indicating that CXCL14 is negatively associated with tumor proliferation (figure 1F, Supple data 2).

To explore the correlation between CXCL14 and immune cells, we utilized the Genotype-Tissue Expression (GTEx) database to analyze the relationship between CXCL14 and various immune cell markers in colon tissue. We observed a strong correlation between the expression level of CXCL14 and T cell markers (CD3, CD4, CD8) in the tissue (r > 0.4, p < 0.001), as well as a robust correlation with the NK cell marker CD122 (r > 0.6, p < 0.001). However, the correlation between CXCL14 and Treg cells, neutrophils, and macrophages is either non-significant or weak (figure 1G, figure S1). This finding suggests that CXCL14 may play a role in tumor immune surveillance by recruiting T cells and NK cells to the colon tissue, aligning with previous research [4].

We also discovered in the Sidra-LUMC tumor database that CXCL14 expression exhibits a significant negative correlation with most matrix metalloproteinases (MMP1, 2, 3, 8, 9, 12, 13, 14, 16, 17, 23b, 25) (figure 1F), indicating that CXCL14 may regulate tumor cell metastasis by controlling the expression of MMP proteins. These findings suggested that CXCL14 is strongly associated with tumor suppressor phenotypes.

### 2. The generation of CXCL14 overexpression cells

CXCL14 is downregulated in human clinical colon cancer samples and tumor cell lines. The mouse colon cancer cell lines MC38 and CT26, as well as the human colon cancer cell line HCT15, are ideal for studying CXCL14 gene overexpression due to their low expression levels. We have successfully generated multiple colon cancer cell lines overexpressing CXCL14. Initially, we utilized the TetOn expression system to establish a tetracycline-inducible expression vector, TetOn-CXCL14, for CXCL14 overexpression (figure S2). Subsequently, MC38 cell line that was stably transfected the plasmid was screened, named MC38-TetOn-CXCL14. This system utilizes doxycycline (DOX)-dependent transcriptional induction to express both murine CXCL14 (mCXCL14) and zsGreen from a bicistronic transcript. The expression of green fluorescent protein serves as a surrogate marker for CXCL14 expression (figure 1B). Flow cytometry analysis revealed that the green fluorescence intensity was comparable to that of the wild-type cells prior to induction but increased by approximately 15-fold following induction (figure S2).

The expression quantity of CXCL14 before and after induction was determined using quantitative PCR, revealing a 12.5-fold increase in the expression level of CXCL14 post-induction. Western blot analysis further supported these findings, indicating a significant increase in the expression level of CXCL14 in cells post-induction (figure1A). We also generated overexpression cell strains of mouse colon cancer cell line CT26 and human colon cancer cell line HCT15 with CXCL14 overexpression.

### 3. Transcriptome analysis of colon cancer cells with CXCL14 overexpression

We utilized doxycycline (DOX) to induce MC38-TetOn-CXCL14 cells to express CXCL14 for 3 days. Cells with and without DOX-induction were collected for RNA sequencing analysis by Geneplus, Beijing, China (see *Data availability statement*). Each sample had two biological replicates: DOX+ cells and DOX-cells. The results of RNA-seq indicated that the expression of the CXCL14 gene was upregulated by a remarkable 14-fold following DOX induction, and WB detection confirmed these findings (figure2A).

The volcanic plot (figure 2B) revealed that following DOX-induced CXCL14 expression, 625 genes demonstrated increased expression while 1237 genes exhibited decreased expression. The GO enrichment analysis showed that the differentially expressed genes were primarily enriched in genes that positively regulate cell motility and migration, angiogenesis, and extracellular matrix-related genes (figure 1C; fold enrichment >2, p <10). These findings suggest that CXCL14 is associated with the regulation of cell invasion ability.

**Figure 2.**
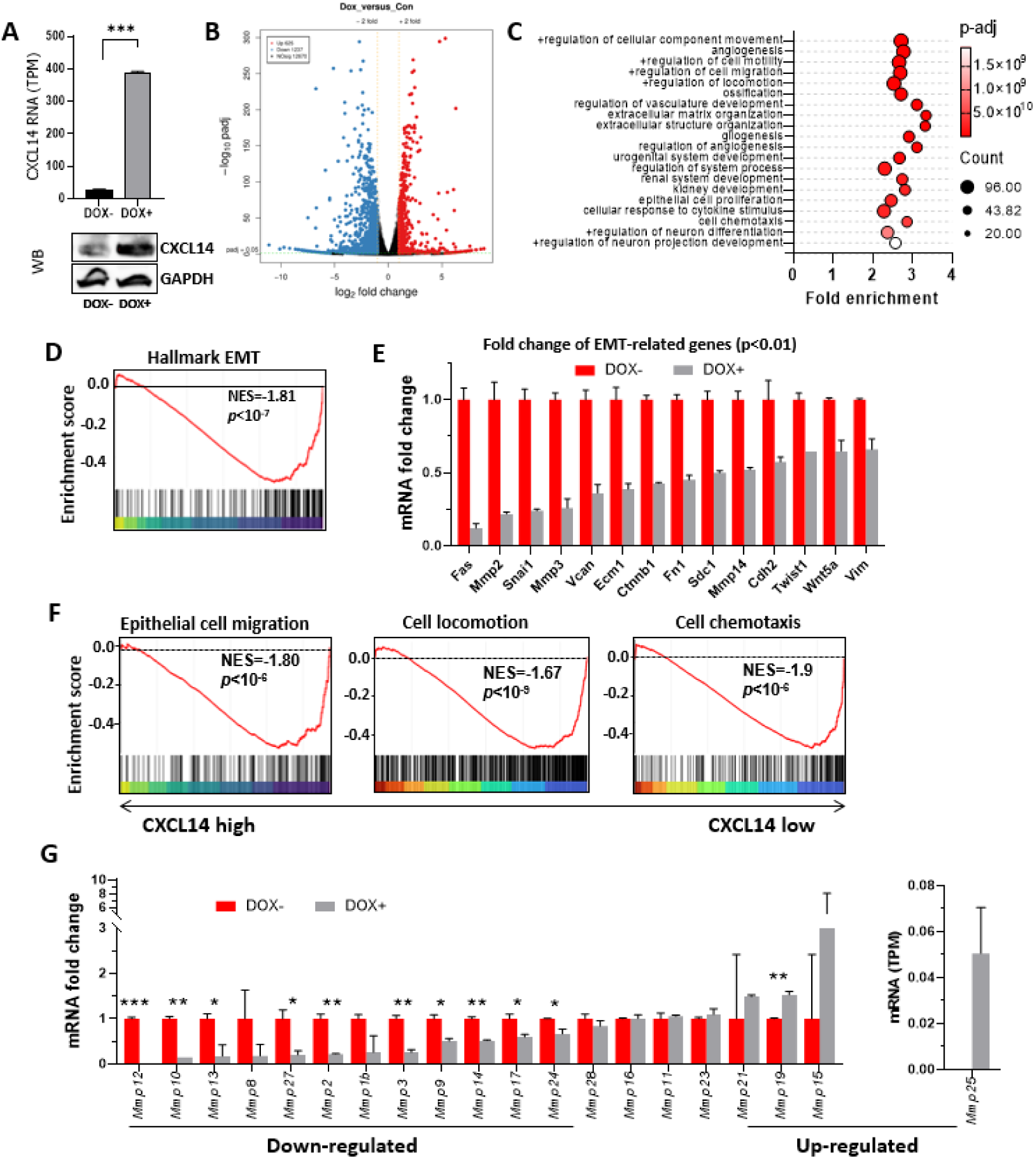
RNA sequencing (RNA-seq) analysis of the regulatory role of CXCL14 overexpression on the transcriptome of MC38 cells. (**A**) RNA-seq data (n=2) and WB result of CXCL14 expression in the non-induced and DOX-induced MC38-TetOn-CXCL14 cell line. **(B)** The volcanic plot displayed that the induction of CXCL14 expression led to a significant upregulation of 625 genes and downregulation of 1237 genes (p-adj < 0.05) in MC38 cells, representing approximately 15% of the total genes. **(C)** The Gene Ontology (GO) analysis of differentially expressed genes (DEGs) between the non-induced and DOX-induced MC38-TetOn-CXCL14 cell line. **(D)** Gene Set Enrichment Analysis (GSEA) enrichment analysis showed that CXCL14 significantly suppressed cell EMT signaling. **(E)** RNA-seq data of EMT-related genes expression of MC38-TetOn-CXCL14 cell line prior or post DOX induction (all p-values < 0.01). **(F)** The GSEA analysis of the impacts of CXCL14 overexpression on MC38 cell migration, locomotion and chemotaxis. **(G)** RNA-seq data of MMP family genes of non-induced and DOX-induced MC38-TetOn-CXCL14 cell line (n=2). Here only genes with non-zero values are displayed. family. *p<0.05, **p<0.01, ***p<0.001.

Using Gene Set Enrichment Analysis (GSEA), we observed that the different expressed genes (DEGs) caused by CXCL14 overexpression were predominantly enriched in the EMT negative regulation-related pathway (figure 2D; NES = –1.81). This suggests that CXCL14 overexpression may impact the EMT characteristics of MC38 cells. We conducted further analysis of the EMT gene set and observed that CXCL14 overexpression led to a significant downregulation of numerous EMT-related genes (figure 2E, p<0.01). These correlations with EMT genes were also confirmed in the tumor database (figure 1E).

The GSEA analysis also revealed that the gene changes caused by CXCL14 overexpression were predominantly associated with the negative regulation of epithelial cell migration, cell motility, and chemotaxis signaling pathways (figure 2E). CXCL14 overexpression was also found to inhibit the expression of numerous genes in the MMPs family (figure 2F). An analysis of the colon cancer Sidra-LUMC AC-ICAM dataset further supported these findings (figure 1H).

The results suggested that CXCL14 may inhibit the development of colon cancer by suppressing tumor cell EMT and cell motility signaling pathways. To confirm this, we searched the TCGA, CPTAC-2, and Sidra-LUMC datasets and found that the expression of CXCL14 is significantly negatively correlated with the stage of colon cancer (figure2G, Supplementary data 3), indicating that the development of colon cancer depends on the silencing of the CXCL14 gene. Clinical pathological sections also showed that CXCL14 expression is significantly reduced in the colorectal tissue (figure2H).

### 4. CXCL14 inhibits the proliferation and motility of colon cancer cells

We have conducted experiments to examine the physiological indicators of cells influenced by CXCL14 overexpression. Notably, CXCL14 overexpression in MC38 cells results in a notable decrease in cell proliferation rate, as clearly demonstrated in figure 3A. To rule out any potential influence of DOX on cell proliferation rate, we evaluated its effect on the growth of wild-type MC38 cells. The results showed that at a concentration of 1 μg/mL, DOX had no discernible impact on MC38 cell proliferation (figure S3A). By expressing CXCL14, the cells exhibited a significant reduction in the expression level of the TK1 gene (as shown in figure3B), which has been established as a crucial marker for tumor cell proliferation. This observation provides further confirmation of the inhibitory effect of CXCL14 on tumor growth, as previously reported in reference [21]. The overexpression of mouse and human CXCL14 genes in CT26 and HCT15 cells has also been observed to inhibit cell proliferation. The intensity of this inhibitory effect is directly proportional to the expression level of CXCL14, as illustrated in figure3C and D. In conjunction with the correlation analysis of TK1 expression levels in the tumor database (figure 1F), it is inferred that CXCL14 may have a role in regulating TK1 expression in colon cancer cells.

**Figure 3.**
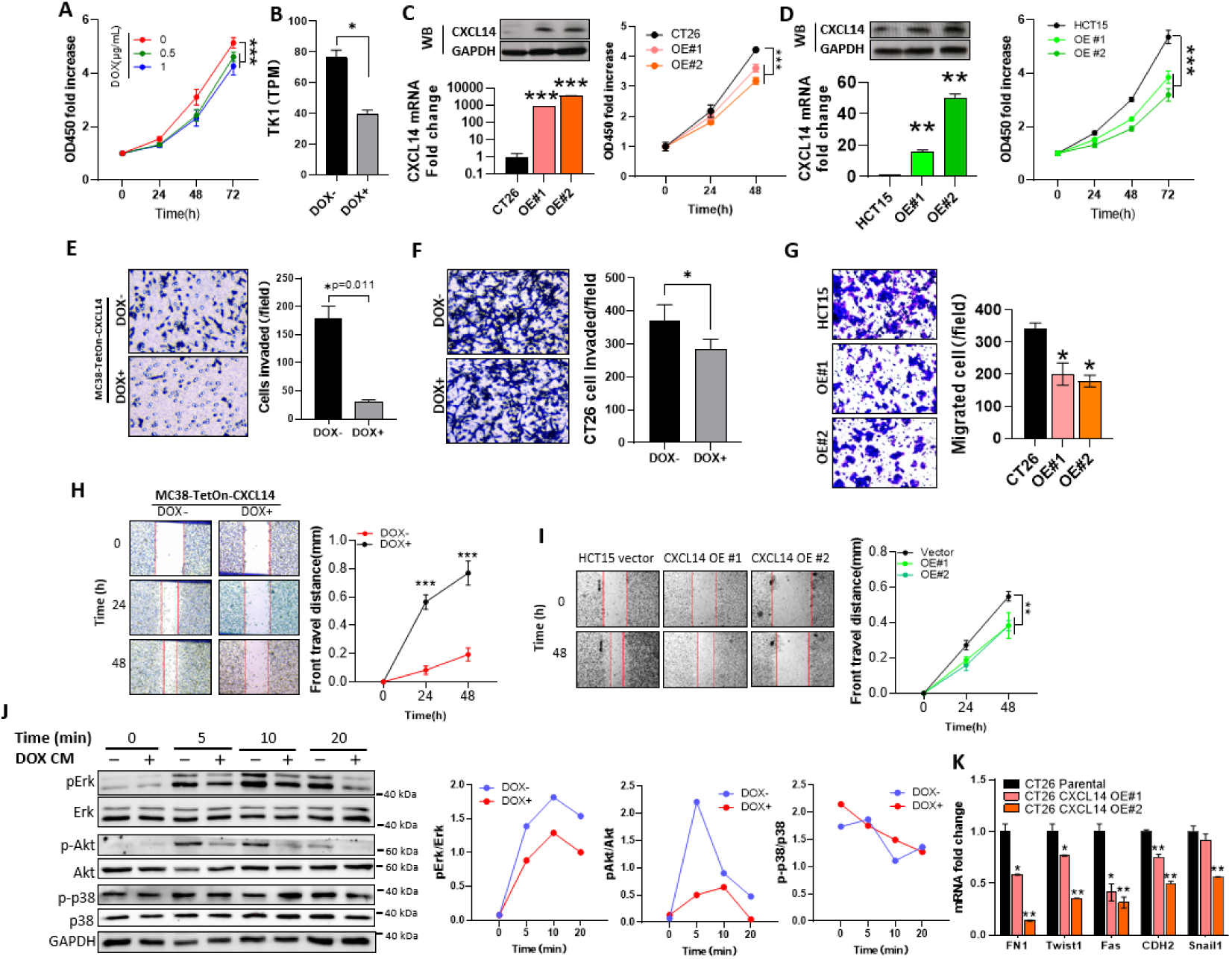
Overexpression of CXCL14 inhibits proliferation, invasion and motility of colon cancer cells. (**A**) Proliferation curves of control and 0.5 and 1 ug/ml DOX-induced MC38 cell (n=6). Data represent the mean±SD. ***p < 0.001, one-way ANOVA. (B) RNA-seq data of TK1 mRNA of control and CXCL14 overexpressed MC38 cell (n=2). Data represent the mean±SD. *p<0.05, t-test. **(C)** WB detection, qPCR data of CXCL14 expression (left) and proliferation curves of parental and CXCL14-overexpressed CT26 cells. Data represent the mean±SD. ***p<0.001, t-test and one-way ANOVA. **(D)** WB assay and qPCR data of CXCL14 expression (left) and proliferation curves of parental and CXCL14-overexpressed HCT15 cells. Data represent the mean±SD. **p<0.01, ***p<0.001, t-test and one-way ANOVA. **(E)** The invasion assay of non-induced and DOX-induced CXCL14-overexpressed MC38 cells (n=3). Data represent the mean±SD. *p=0.011, t-test. **(F)** The invasion assay demonstrates that overexpression of CXCL14 in CT26 cells impairs their invasion capability (n=3), *p<0.05, t-test. **(G)** The invasion assay has revealed that the restored expression of CXCL14 in HCT15 cells effectively suppresses their invasion capacity. **(H)** The wound repair assay of control and 1 ug/ml DOX-induced MC38 cell (n=10). Data represent the mean±SD. ***p < 0.001, t-test. **(I)** The wound repair assay of parental and CXCL14-overexpressed HCT15 cell lines (n=6). Data represent the mean±SD. ***p < 0.001, one-way ANOVA. **(J)** WB assay of the effect of CXCL14-conditioned medium on the phosphorylation of Erk, Akt, and p38 proteins in MC38 cells. The left panel of the figure displays the WB results, and the right panel represents the consistent outcomes from multiple replicates of the experiment. **(K)** qPCR data of EMT-related genes in parental and CXCL14-overexpressed CT26 cells (n=2), *p<0.05t-test.

CXCL14 exerts a significant impact on cell invasion. When the MC38-plvx-TetOn-CXCL14 cells were induced to express CXCL14 with doxycycline, their invasion capacity was significantly reduced (figure 3E). To rule out the possibility that DOX was responsible for changes in invasion function, we compared invasion capacity of wild-type MC38 cells grown with and without DOX (1 μg/mL). figure S3B shows that DOX did not inhibit invasion ability of MC38 cells. We transiently transfected MC38 cells with CXCL14 and observed a significant reduction in their invasion capacity (figure S4). This inhibitory effect on invasion was also observed in CT26 and HCT15 cells (figure 3F, G).

Using the scratch test, we evaluated the impact of CXCL14 on cell motility and observed a significant reduction in the motility of mouse colon cancer MC38 cells and human colon cancer HCT15 cells (figure 3H, I). Furthermore, CXCL14 exerted a significant inhibitory effect on key EMT genes in HCT15 cells (figure S5).

Chemokines generally regulate the physiological state of cells through the GPCR-mediated MAPK signaling pathway. Therefore, we employed CXCL14-conditioned medium to investigate its regulatory effect on the MAPK signaling pathway in MC38 cells. We collected cell lysates at 5, 10, and 20 minutes post-medium addition and evaluated the phosphorylation levels of MAPK-related kinases. The results indicated that compared to the wild-type MC38 medium, the conditioned medium of CXCL14 overexpressing cells significantly inhibited the phosphorylation of Erk and Akt, but had no significant effect on p38 phosphorylation (figure 3J).

We tested the EMT-related genes expression with CXCL14 overexpression in CT26 cells. The results of quantitative PCR indicate that overexpression of CXCL14 has a suppressive impact on the expression of CT26 EMT-related genes (figure 3K). Furthermore, the magnitude of this inhibitory effect is directly proportional to the extent of CXCL14 overexpression, as illustrated in figure 3C.

### 5. CXCL14 inhibits the proliferation of subcutaneous tumor burden

To regulate the overexpression of CXCL14 in subcutaneously inoculated MC38 cells, we employed the Tet-inducible expression system. The experiment consisted of two groups of animals, the induced group and the non-induced group, with 8 animals in each group. The experimental procedures are described in “Method 3”. After overexpression of CXCL14, the tumor growth rate was significantly reduced (p<0.001, figure 4A). At the end of the experiment, the tumor size and weight in the CXCL14 overexpression group was significantly smaller than that in the control group (figure 4B, C). Throughout the entire experimental period, there was no significant difference in body weight between the two groups of mice (figure4D).

**Figure 4.**
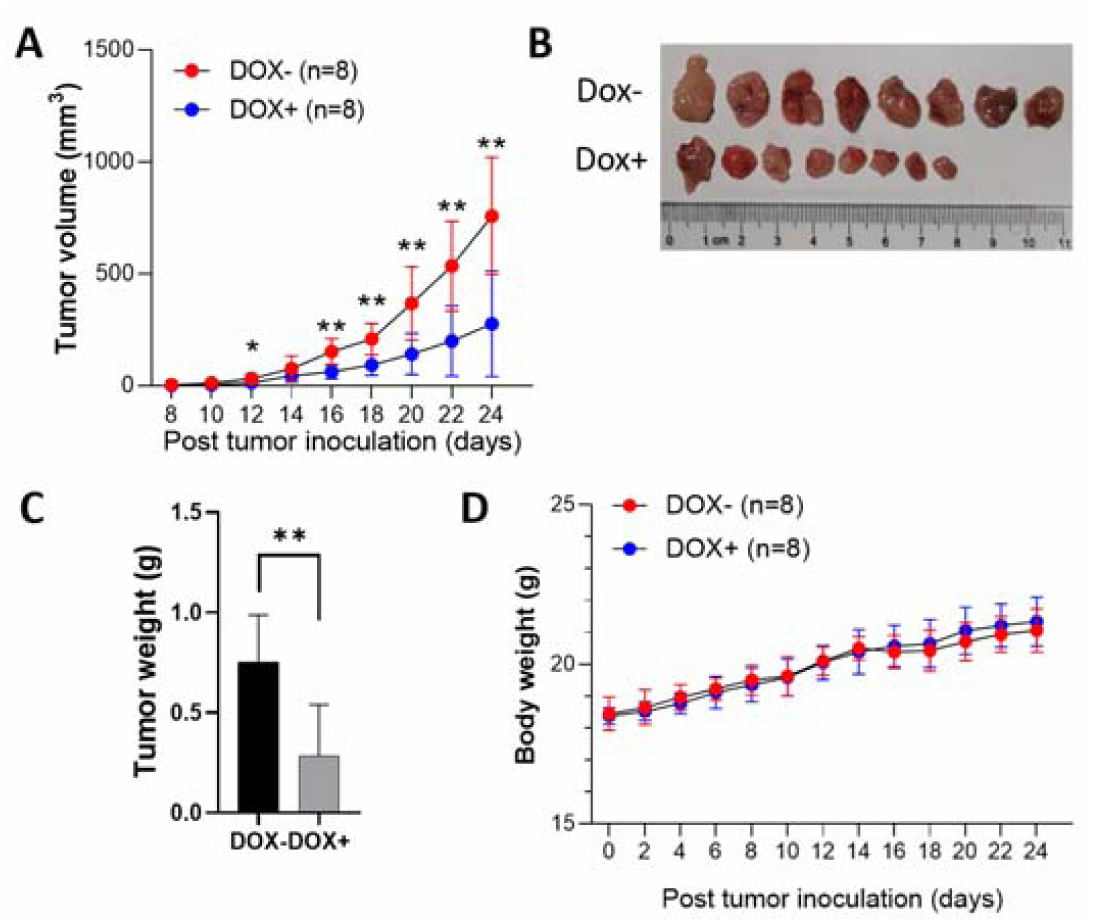
The overexpression of CXCL14 hinders the proliferation of MC38 cells in the subcutaneous tissue of mice. (**A**) The tumor growth curves of DOX-induced and non-induced groups of C57bl/6n mice with subcutaneous MC38-plvx-TetOn-CXCL14 cells. The tumor volume of the DOX-induced group was significantly lower than that of the non-induced group (n=8), *p<0.05, **p<0.01, t-test. **(B)** Images of tumors from both groups of mice (n=8). **(C)** The tumor tissue was excised from the subcutaneous tissue and weighed. The tumor weight of the DOX-induced group was significantly lower than that of the non-induced group (n=8), p<0.01, t-test. **(D)** The body weight curves of non-induced and DOX-induced mice (n=8), there was not significant difference by t-test.

### 6. CXCL14 regulates tumor immune microenvironment

We used MC38-TetOn-CXCL14 cells to induce subcutaneous tumors in C57BL/21 mice. Once the tumor volume reached 1000 mm^3^ , the mice were administered with drinking water containing 1 mg/mL DOX. Frozen sections of the tumor tissue revealed significant expression of the reporter gene EGFP (figure 5A). RNA-seq analysis (see *Data availability statement*) revealed a significant increase in the expression level of CXCL14 in the DOX-induced group (figure 5B). The volcano plot showed that CXCL14 induction led to an increase in the expression of 720 genes and a significant decrease in the expression of 234 genes, representing 5.7% of the total genes detected (figure 5C).

**Figure 5.**
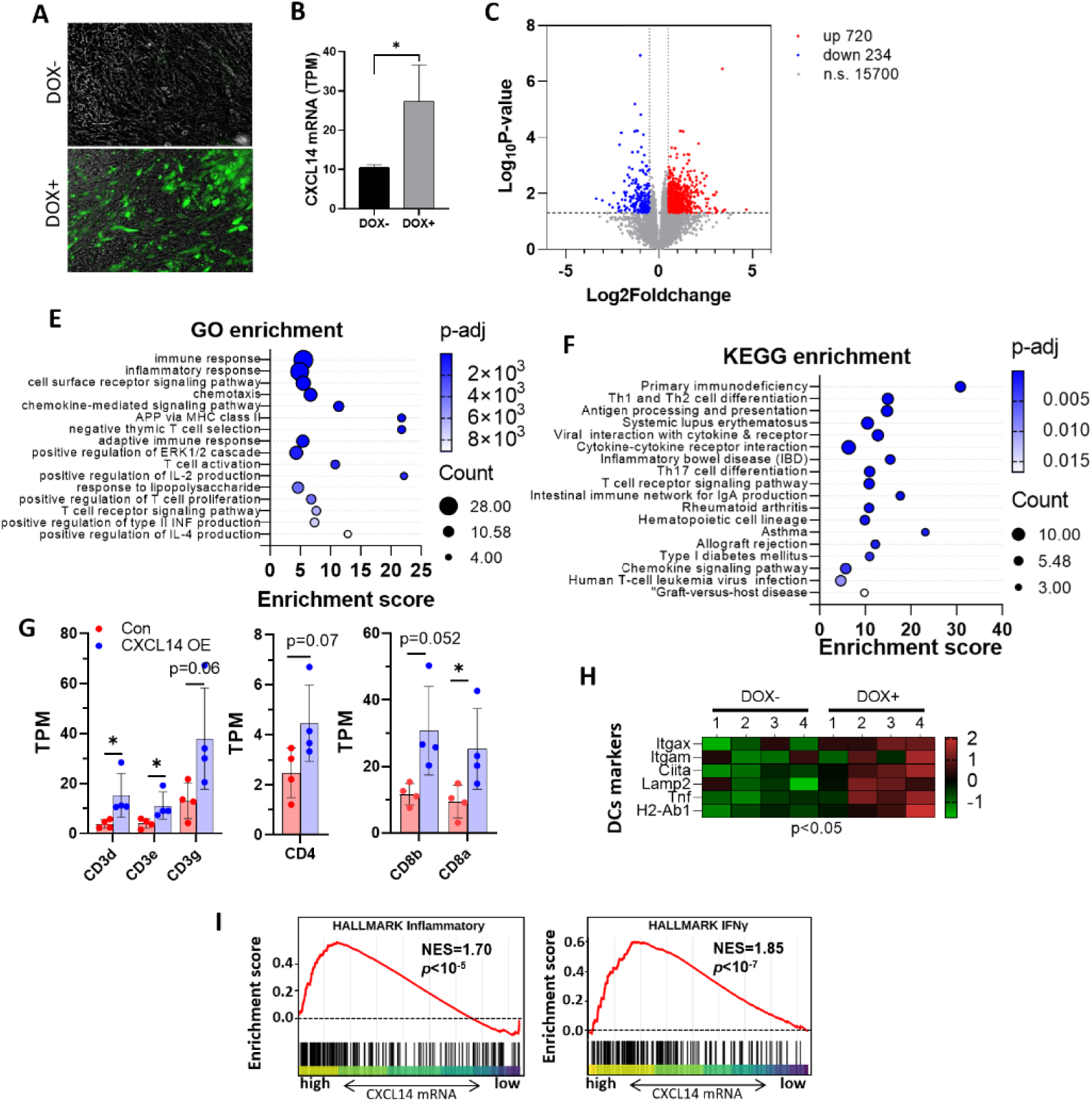
Transcriptome analysis showed that CXCL14 regulates the tumor microenvironment. (**A**) Frozen sections from mice bearing subcutaneous tumors revealed the presence of the reporter gene EGFP, indicating the induction of CXCL14 expression within the tumor. (B) RNA-seq data of the expression level of CXCL14 in non-induced and DOX-induced CXCL14 overexpression tumor tissues (n=4), *p<0.05, t-test. (C) RNA-seq results revealed that there were 720 significantly up-regulated genes and 234 down-regulated genes in CXCL14 OE MC38 subcutaneous tumors compared to MC38 non-induced tumors. (D) The GO analysis results indicate that the gene changes caused by CXCL14 overexpression are mainly concentrated in immune/inflammatory response, chemotaxis, and T cell-related signal transduction. The GO enrichment results obtained in this experiment were compared with the signal transduction obtained by enriching the top 1000 genes related to CXCL14 in the Gepia database. It was found that there were 16 overlapping terms. The KEGG enrichment analysis results indicate that the gene changes caused by CXCL14 overexpression are mainly concentrated in immune deficiency, Th cell differentiation, antigen presentation. Compared with the clustering results of the top 1000 genes related to CXCL14 expression in the Gepia database, it was found that the relevant terms highly overlap (21 overlapping terms). (E) The 16 overlapping signal transduction pathways in figure D are mainly concentrated in immune response, chemotaxis, antigen presentation, and T cell activity regulation. **(F)** The KEGG pathway enrichment analysis in figure D reveals that there are 21 overlapping signal transduction pathways between the two datasets, including T cell differentiation, antigen presentation, and immune enteritis. **(G)** The RNAseq data of T cell marker genes in the subcutaneous tumor with non-induced and DOX-induced CXCL14 overexpression (n=4), *p<0.05, t-test. **(H)** The heat map of qPCR data of dendritic cell markers in the subcutaneous tumor with non-induced and DOX-induced CXCL14 overexpression (n=4), p<0.05, t-test. **(I)** GSEA of hallmark inflammatory and IFNγ gene signatures was performed to compare CXCL14 overexpressed MC38 cells with non-induced MC38 cells.

To validate the consistency of CXCL14 function in mouse models and human colon, we used the Gepia website (http://gepia2.cancer-pku.cn) to retrieve the 1000 genes with the strongest correlation with CXCL14 expression in colon tissue from the GTEx database (Supplementary data 4). Subsequently, the GO and KEGG clustering analysis were performed on these 1000 genes, and the terms with p-value (adjusted) <0.01 were screened out, obtaining 77 GO terms and 31 KEGG terms, respectively (Supplementary data 5 and 6). These clustering results were compared with the differential gene clustering results of subcutaneous tumor-bearing mice, and it was found that there were 16 overlapping terms in the GO clustering results (figure5E), while there were 18 overlapping terms in the KEGG clustering results (figure5F). The above results indicate that the function of CXC14 gene in MC38 subcutaneous tumor-bearing mice is highly consistent with that in human colon tissue.

The GO cluster analysis showed that CXCL14 affects a large number of signal pathways related to immunity, including immune response, inflammatory response, adaptive immune response, T cell activation, IFNγ, IL2, IL4 production, etc. (figure5E). The regulatory network generated by the enrichment analysis is shown in figureS6. The KEGG cluster analysis reflects the role of CXCL14 in the tumor microenvironment from another perspective, mainly focusing on the effect of T cells, including Th1, 2, 17 cell differentiation, antigen presentation, etc. (figure5F). The RNA-seq results showed that the levels of T cell markers and surface markers of Tc and Th cells were significantly higher than those of the control group (figure5G). A series of DC cell markers were also significantly elevated in the tumor overexpressing CXCL14 (figure5H). We also performed GSEA with the RNA-seq data from CXCL14-overexpressed and uninduced cells and demonstrated significant enrichment of the gene signatures related to inflammatory pathway and IFNγ production (figure4I).

### 7. CXCL14 mediates T-cell infiltration into subcutaneous tumors

We conducted histological studies on DOX-induced subcutaneous tumors and observed that the tumor tissues overexpressing CXCL14 had more necrotic tissue areas (figure 6A). Immunostaining with the CD3 antibody revealed a significant increase in the number of CD3+ positive cells (figure 6B). Flow cytometry analysis of total tumor cells revealed that T cells accounted for approximately 1% of the total cells in the tumor tissues (figure 6D). Analysis of CD4 and CD8 cell numbers (figure 6E) showed that the proportion of CD4 T cells was significantly increased in the tumor overexpressing CXCL14, while the proportion of CD8 T cells was slightly increased (figure 6F, G). The flow cytometry analysis of macrophage levels revealed that there was no significant increase in the count of CD40 and CD11b double-positive macrophages.

**Figure 6.**
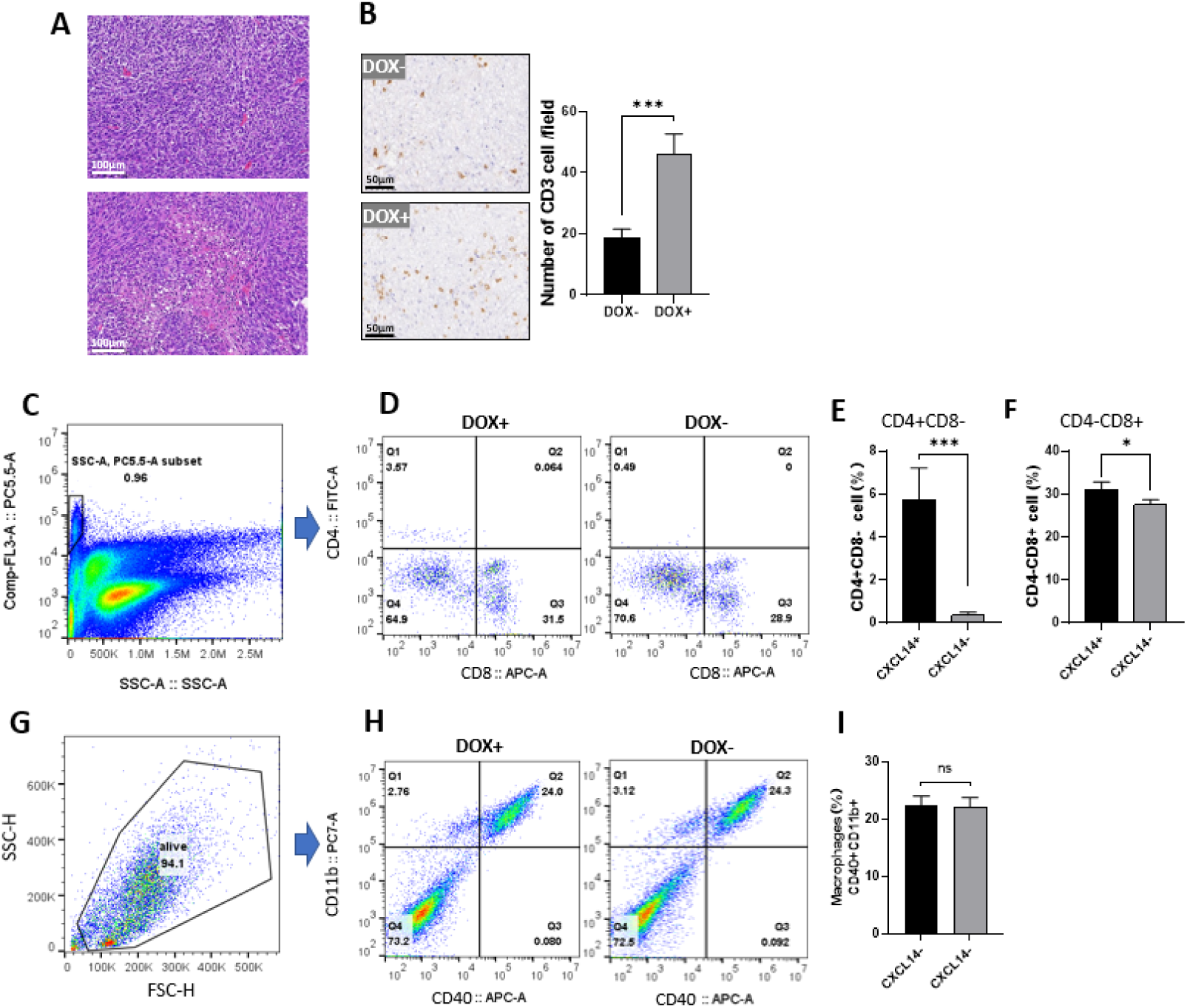
CXCL14 enhances tumor T cell infiltration. (**A**) The H-E staining of the tumor showed that there were more necrotic areas in the tumor overexpressing CXCL14. (**B**) The abundance of CD3 cells was significantly increased in the tumor tissues induced by DOX+ overexpression of CXCL14. Each treatment had 4 biological replicates, and 5 high-power fields were randomly selected from each tumor to count the positive cells. (**C**) The CD3 (PE-Cy5.5) and SSC scatter plot of the tumor cell suspension. The gate in the figure selects CD3-positive T lymphocytes. (**D**) The flow cytometry results of CD4 (FITC) vs CD8 (APC). The Q1 quadrant represents CD4+CD8-T lymphocytes, and the Q3 quadrant represents CD4-CD8+T lymphocytes. (**E**) The statistical results of the Q1 quadrant cells in figure D showed that CD4+CD8-T lymphocytes were significantly increased in the tumor cells induced by DOX overexpression (p<0.001). (**F**) The statistical results of the Q3 quadrant cells in figure D showed that CD4-CD8+ T lymphocytes were significantly increased in the tumor cells induced by DOX overexpression (n=4), *p<0.05, ***p<0.001, t-test. (**G**) The scatter plot of FSC/SSC of the tumor cell suspension. The gate encloses live cells. (**H**) The scatter plot of CD40/APC and CD11b/PC7 of tumor cells. The Q2 quadrant represents double-positive cells, which are tumor macrophages. (**I**) There was no significant change in the content of macrophages in tumor-infiltrating cells(n=4), t-test.

## Discussion

CXCL14 is one of the most highly conserved members within the chemokine family, as demonstrated by Huising et al [9]. It originated from two gene duplications of the CXCL ancestor gene, and has since undergone independent evolution. The Ks/Ka ratio highlights that CXCL14 is subject to the most stringent purifying selection pressure among all CXC chemokines, as depicted in figureS7 [22, 23]. This further underscores its critical role in maintaining its functions.

Despite its significant role, CXCL14 remains enigmatic within the chemokine family. The receptor for this molecule remains unresolved, with ongoing debate centering on whether CXCR4 serves as its natural receptor [12–15]. Using the b-Arrestin Assay, Kouzeli et al. conducted a comprehensive investigation into the interactions of CXCL14 with known chemokine receptors and found that CXCL14 is unable to activate any members of the chemokine receptor family, including CXCR4, independently. Instead, it solely participates in signal transduction functions in conjunction with other ligands through synergistic interactions [17].

Whether there are receptors outside the chemokine ligand family remains uncertain. However, a recent study suggests that MRGPRX2 may serve as the natural receptor for CXCL14 [16]. Nevertheless, there is a contradiction in that CXCL14 is a highly conserved gene (95% identity between human and mouse proteins), whereas MRGPRX2 is not (54% identity). Furthermore, MRGPRX2 is not expressed on immune cells, thus even if MRGPRX2 is the receptor for CXCL14, there could still be another receptor responsible for mediating the chemotactic effect of CXCL14 on immune cells. A study exploring the interactions among chemokine ligands has revealed extensive heterodimerization among members of the chemokine family. This heterodimerization transforms the previously two-dimensional interaction network of ligands and receptors into a more complex three-dimensional network[18]. When coupled with the intricate dimerization and polymerization phenomenon observed among chemokine receptors, it becomes evident that the system is significantly more complex than what was previously envisioned [24].

Chemokines play a pivotal role in modifying the tumor microenvironment. However, given the diverse nature of chemokines as outlined above, the same chemokine ligand can exhibit distinct microenvironment regulatory functions and corresponding prognosis in different tumor types. CXCL14 serves as a prime example of a multifunctional factor. Shabgah et al. and Westrich et al. have reviewed the distinct expression change trends and prognosis of CXCL14 in various organ tumors [25, 26].

In colorectal cancer, CXCL14 exhibits a more tumor-suppressing effect. Previous studies have shown that CXCL14 can inhibit tumor proliferation, migration, and invasion, among other functions, at an *in vitro* level[6, 7]. Additionally bioinformatics research has also revealed the phenomenon of methylation-induced silencing of CXCL14 in colon cancer, which is associated with poor prognosis [5]. This supports the hypothesis that CXCL14 acts as a tumor suppressor gene in colon cancer.

In this study, we have shown that CXCL14 regulates the EMT characteristics of tumor cells. EMT, a cell migration mechanism during embryonic development, can also activate tumor cells’ abilities to proliferate and invade[27]. Inducing the reverse process of MET has been proposed as a potential new treatment strategy for tumors. CXCL14, one of the earliest chemokines, is thought to play a role in neural cell migration[9]. Its knockout can lead to partial embryonic death, highlighting the crucial role of CXCL14 in embryonic development[28]. From an evolutionary perspective, the emergence of CXCL14 aligns with the development of the neural system in the course of biological evolution[9].

We have demonstrated through gene knockout mice that CXCL14 has an important impact on neural migration in the cerebral cortex and embryonic survival rate [29]. Therefore, we believed that CXCL14 is an important gene that regulates EMT in colon cancer, and CXCL14 gene silencing is one of the switches for tumor cells to enter the EMT stage.

Some data from this study provide support for the aforementioned view. Firstly, the overexpression of CXCL14 resulted in a downregulation of 12 MMP members, which are crucial markers of EMT (figure2G). Additionally, Kuang et al. reported that CXCL14 negatively regulates MMP2 and MMP9 expression in trophoblast cells [30]. Secondly, through analysis of multiple clinical colon cancer databases, we found that CXCL14 is negatively associated with key genes involved in EMT, including FN1, CDH2, and most metalloproteinases (figure1E,1H). These findings suggest that CXCL14 may inhibit tumor metastasis and invasion by promoting the MET process.

Angiogenesis is an important mechanism for suppressing tumor growth. According to the results of transcriptome differential enrichment analysis, CXCL14 can regulate the expression of angiogenesis-related genes. Among them, ADGRG1, an angiogenesis inhibitor gene[31, 32], was strongly up-regulated by CXCL14 in both in vitro and in vivo models in this experiment (figureS8), which deserves further attention.

When it comes to signal transduction, we have evaluated the impact of CXCL14 on the MAPK signaling pathway, a canonical GPCR-mediated intracellular pathway that regulates cell motility, growth, and differentiation. Our experimental findings demonstrate a notable inhibitory effect of CXCL14 on MAPK (see figure 3K).

In terms of signal transduction, we have tested the effect of CXCL14 on the MAPK signaling pathway, which is a typical GPCR-mediated intracellular signaling pathway that affects cell motility, growth, and differentiation. The experimental results show that CXCL14 has a significant inhibitory effect on MAPK (figure3K). Nevertheless, the precise signal transduction mechanism remains to be further investigated.

Previous studies have demonstrated that CXCL14 regulates the tumor microenvironment by influencing DC cell infiltration in human head and neck cancer and DC cell chemotaxis in vitro [1, 33], as well as recruiting NK cells in the uterus of mice and NK cells in vitro[4, 34]. This study found that CXCL14 not only recruits DC and NK cells, but also has the function of recruiting and activating T cells, which was first discovered in colon cancer. Parikh et al. found that overexpression of CXCL14 in oral cavity squamous cell carcinoma can recruit T cell infiltration, and deletion of T cells eliminates the tumor-suppressing effect of CXCL14 [35], which is consistent with our findings in this study.

Some of the phenomena revealed in this study are worthy of further investigation. Among them, the conditioned medium of CXCL14 can significantly inhibit the AKT and ERK signaling pathways. However, whether CXCL14 directly acts on a receptor or whether the downstream cytokines activated by the CXCL14 signaling pathway lead to this result needs to be studied in detail.

## Conclusion

In summary, our study demonstrates that CXCL14 exerts an inhibitory effect on the development of colon cancer, which is attributed to two functional aspects. Initially, CXCL14 exhibits the capacity to suppress the proliferation, migration, and metastasis of tumor cells. Secondly, CXCL14 also modulates the immune microenvironment of colon cancer, increasing the levels of NK, DC, and T cells, and converting the tumor from a “cold” to a “hot” phenotype. These findings suggested that CXCL14 could serve as a potential target for immunotherapy in colon cancer.

## Data availability statement

Data are available in a public, open access repository. The MC38 cell RNA-seq data are available through GEO access number GSE256328. The Subcutaneous tumor RNA-seq data are available through GEO access number GSE256330.

## Supplementary Figures

**Supplementary Figure 1.**
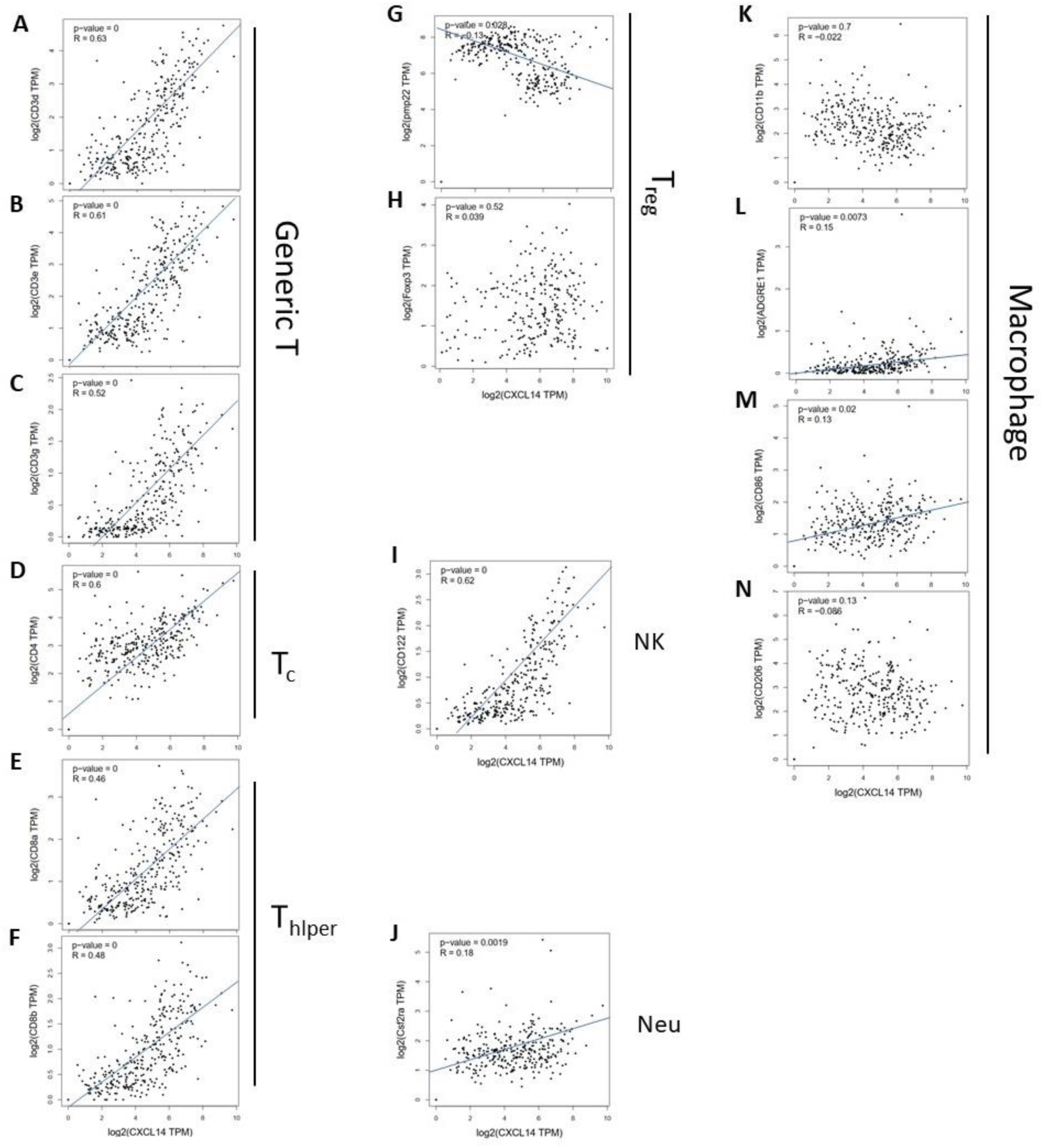
The correlation coefficients of CXCL14 and immune cell biomarkers. (A-C). CXCL14 in colon tissue (Sigmoid and Transverse) is expressed strongly correlated (p<10^8^, R>0.5) with T cell marker CD3d, CD3e, and CD3f; (D). CXCL14 in colon tissue (Sigmoid and Transverse) is expressed correlated with Tc biomarker CD4 (p<10^8^, R>0.4); (E-F). CXCL14 expression in colon tissue is strongly correlated with T help biomarker CD8a, CD8b(p<10^8^, R>0.4); (G-H). CXCL14 shows no or negative correlation with Treg biomarker Pmp22 and Foxp3. (I). CXCL14 shows significant positive correlation with NK biomarker CD122. (J). CXCL14 shows weak positive correlation with Neu biomarker Csf2ra (R=0.18). (K-N). CXCL14 shows weak or no correlation with Macrophage biomarker CD11b, ADGRE1(R=0.15), CD86(R=0.13) and CD206

**Supplementary Figure 2.**
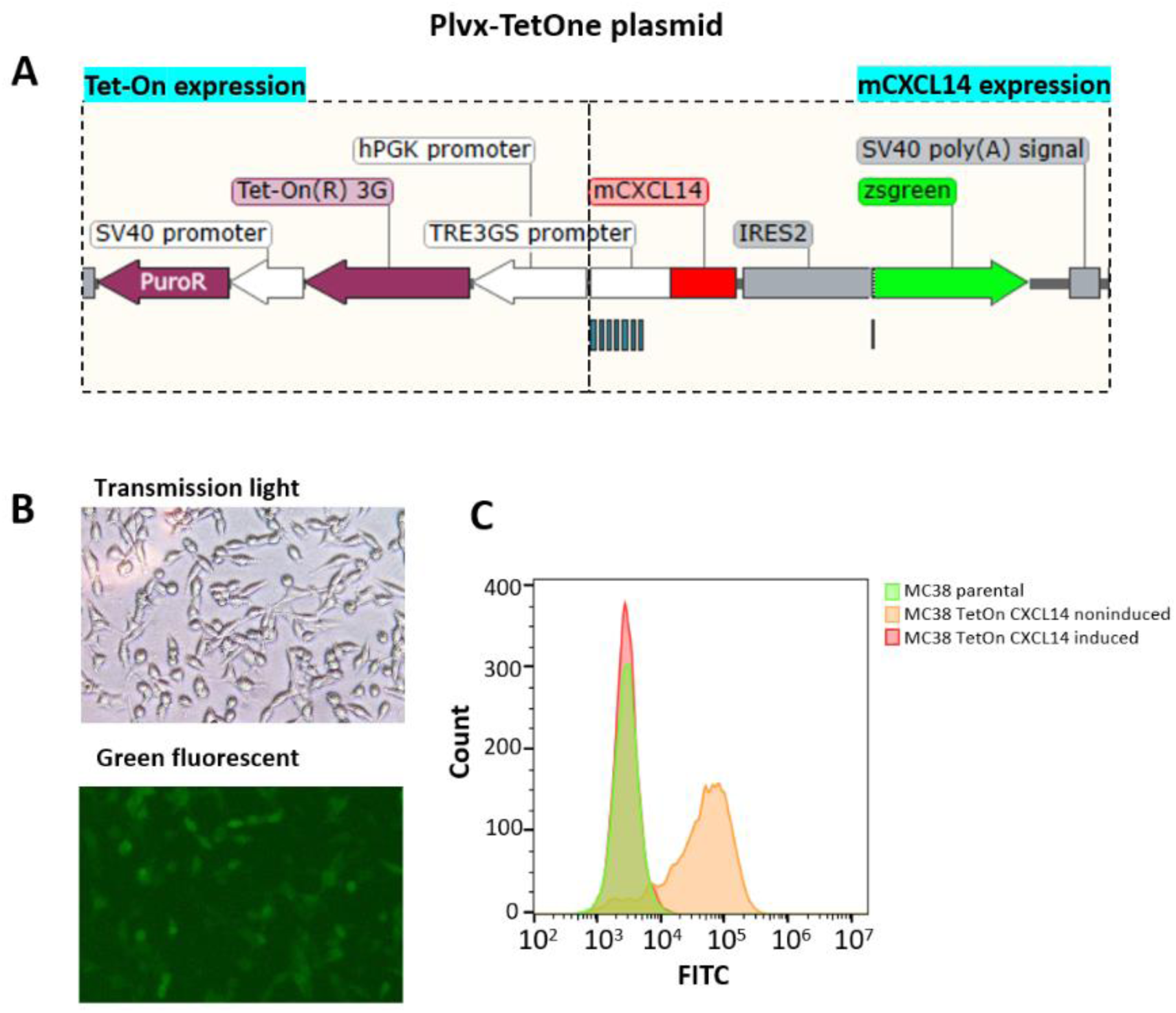
Structure of CXCL14 inducible plasmid and expression. (A) The structure of the inducible CXCL14 expression plasmid. The plasmid is derived from the tetracycline-inducible expression plasmid system. The plasmid constitutively expresses the Tet-On(R)3G protein, which has promoter activity in the presence of tetracycline derivatives and can bind to the TRE3GS promoter to activate the mRNA transcription of downstream genes. The transcriptional unit in the plasmid includes the mouse CXCL14 gene, the IRES2 sequence, and the zsgreen gene. The zsgreen gene serves as a fluorescent reporter gene to indicate the expression of mCXCL14 gene. (B) After inducing MC38-TetOn-CXCL14 cells with 1ug/mL DOX for 48h, fluorescence microscopy images indicate that the reporter gene zsgreen is clearly expressed in the cells. (C) Flow cytometry was used to detect the expression of EGFP in MC38-TetOn-CXCL14 cells before and after induction. The cells before induction were similar to parental MC38 cells, while the signal was significantly increased after induction.

**Supplementary Figure 3.**
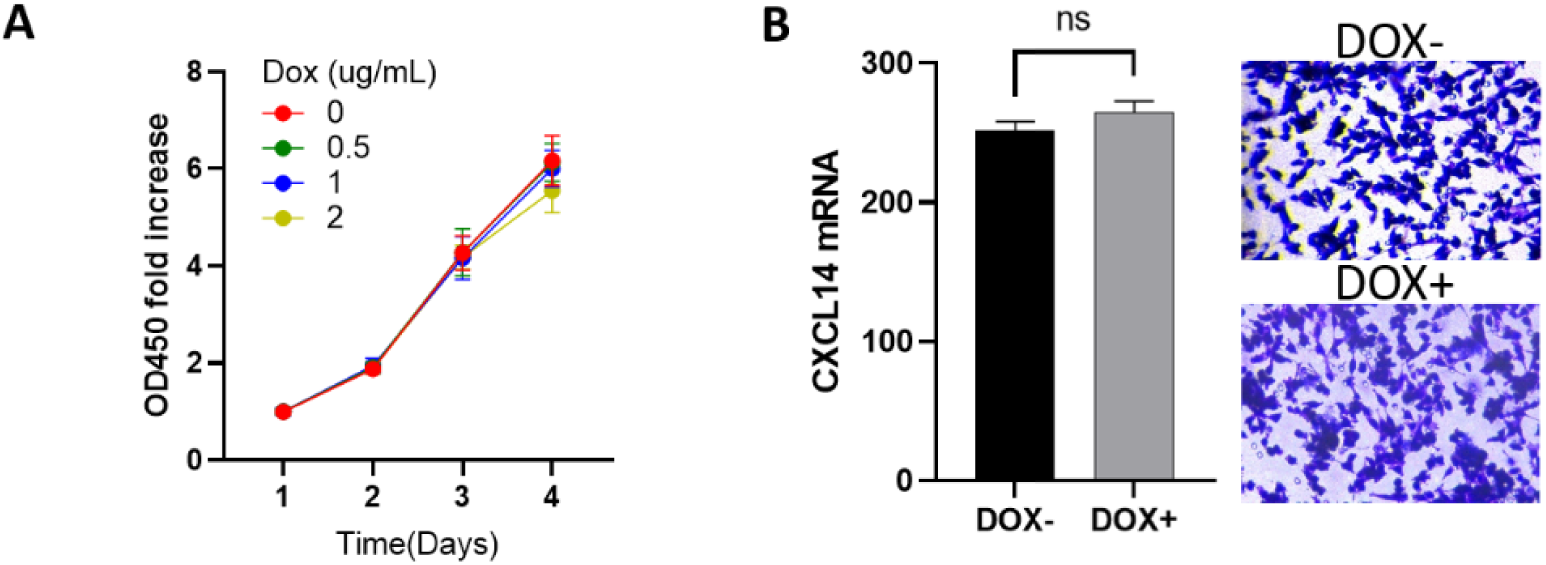
DOX does not affect the proliferation and invasion of MC38 cells. (A) The growth rate of MC38 cells cultured with 0.5∼2ug/ml doxycycline is not affected. (B) The tumor invasion ability of MC38 cells cultured with 1 ug/ml doxycycline is not significantly changed.

**Supplementary Figure 4.**
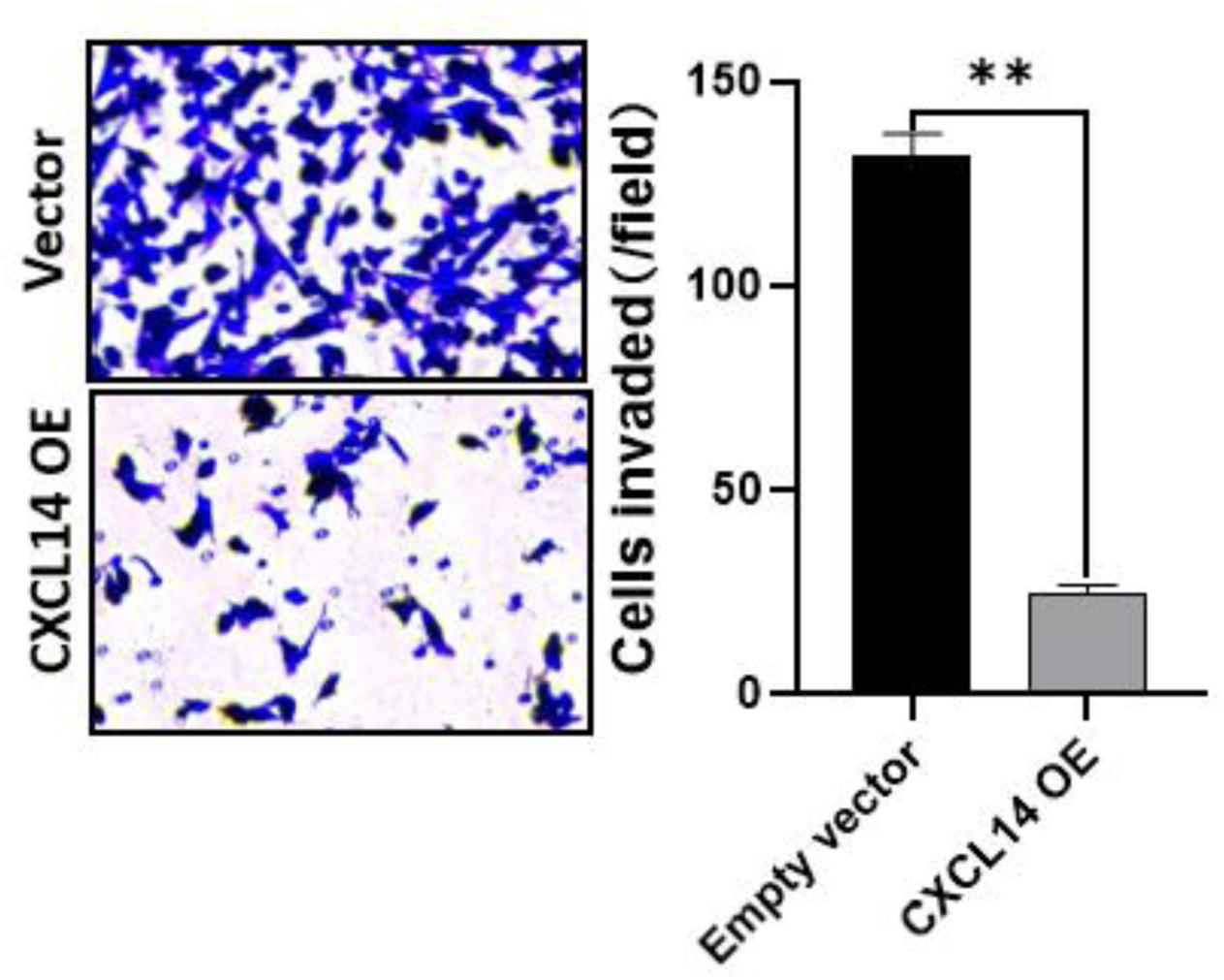
Transient transfection of CXCL14 inhibit MC38 cell invasion. The transient transfection of MC38 cells with the plasmid pcDNA-CXCL14 for overexpression of CXCL14 can significantly inhibit the invasive ability of the cells (n=3). **p<0.01, t-test.

**Supplementary Figure 5.**
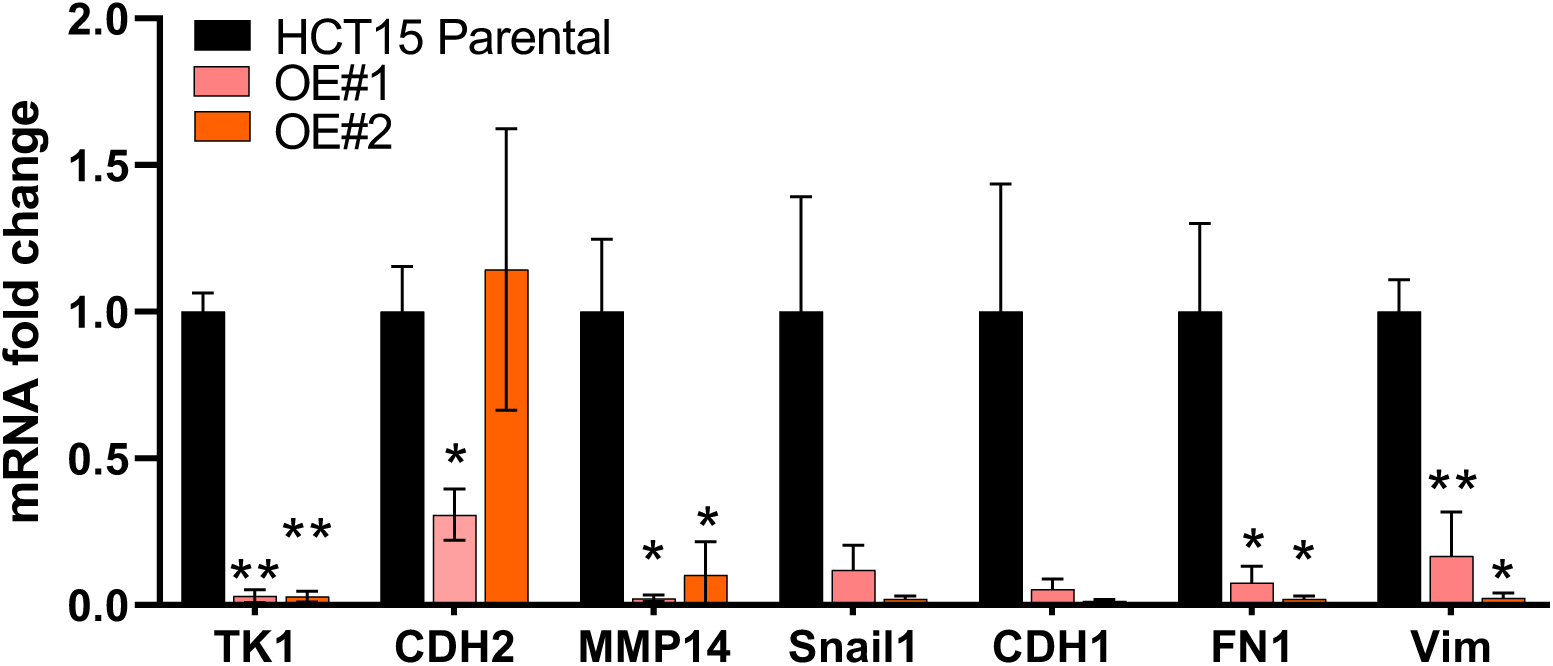
The overexpression of CXCL14 inhibits the expression of EMT-related genes in HCT cells. Q-PCR analysis of the expression levels of pertinent genes in HCT15 cell lines overexpressing CXCL14, it was observed that the overexpression of CXCL14 led to significant inhibition of tumor proliferation marker (TK1) and tumor EMT markers (n=2). *p<0.05, **p<0.01, t-test.

**Supplementary Figure 6.**
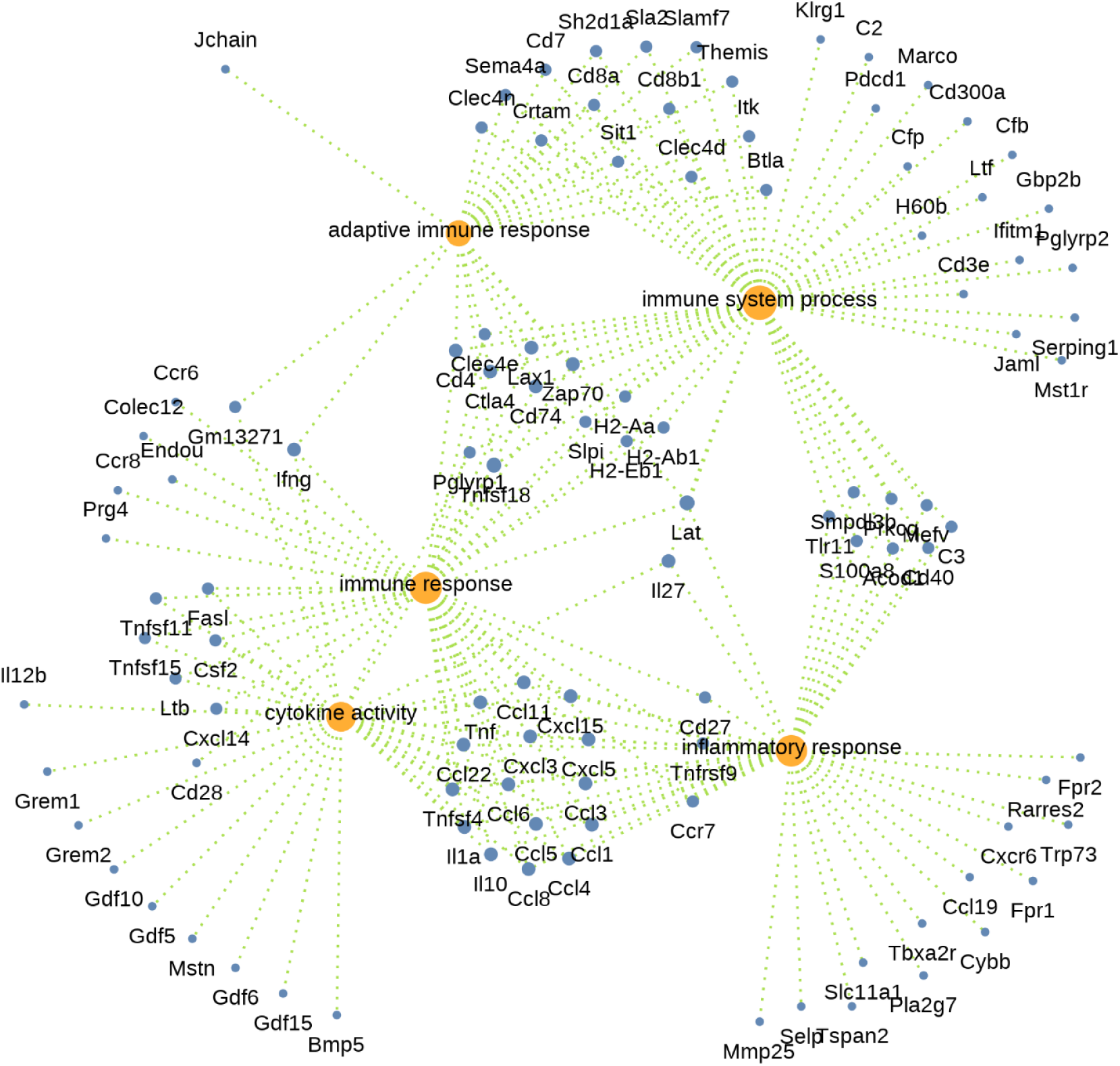
GO-dotplot of DEGs in subcutaneous tumors. The DEGs between the non-induced and DOX-induced MC38-TetOn-CXCL14 tumor were analyzed by GO.

**Supplementary Figure 7.**
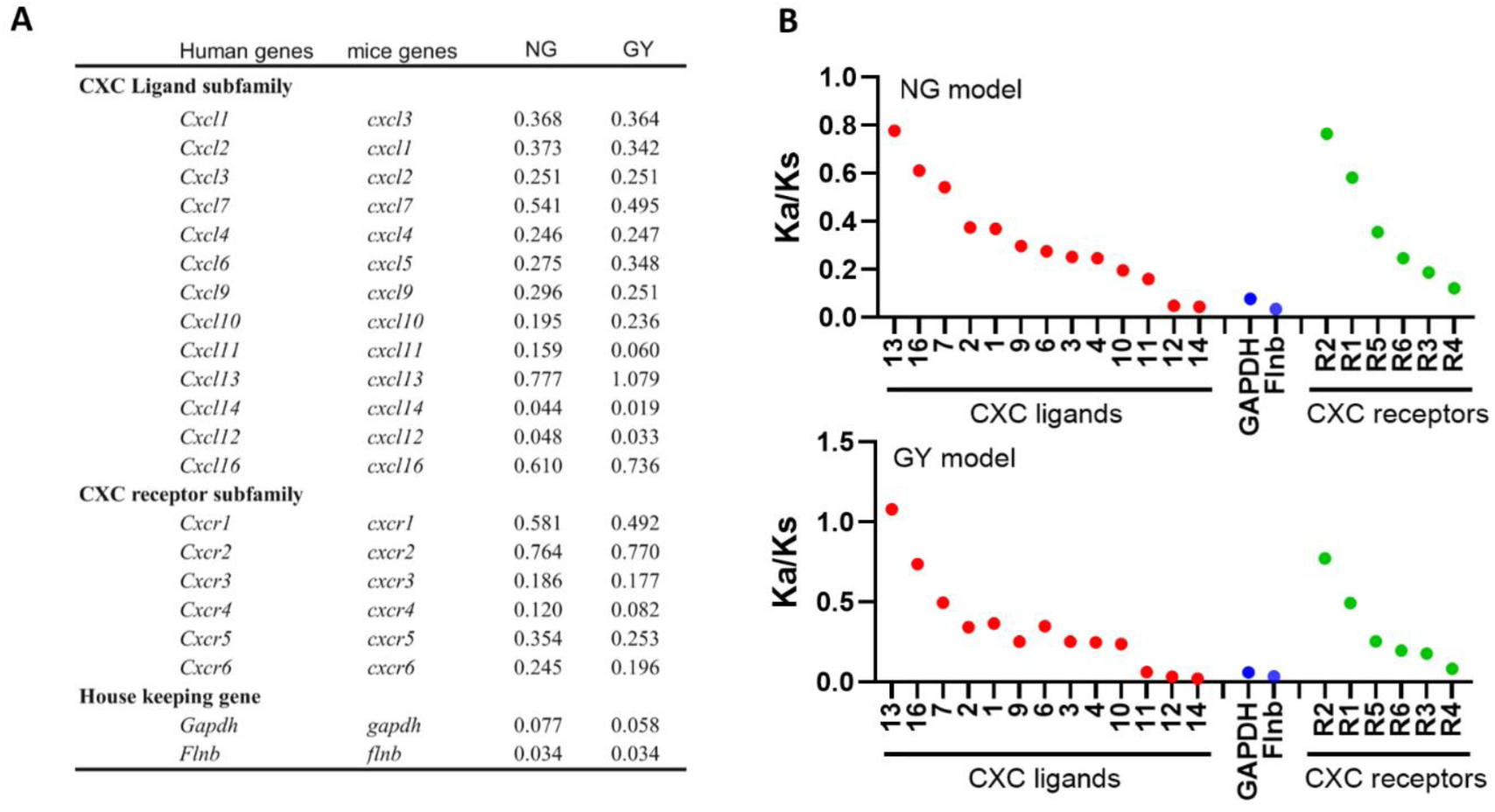
Ka/Ks ratio of CXC chemokine and receptor family. (A) The left column shows the Ka/Ks of each gene calculated using software KaKs_calculator Version 1.2 (http://evolution.genomics.org.cn/software.htm) with homologous genes of human and mouse by NG, an approximate algorithms [122], and GY, a Maximum-Likelihood Methods (Goldman N., et al. Mol Biol Evol, 1994, 11: 725-736). Ka/Ks is calculated in mature peptide of chemokines, ectodomains of receptors, and *Gapdh* and *Flnb* of human and mouse as references. **(B)** Scatter diagram of Ka/Ks of ligand and receptor family. The results of two algorithems insistently indicated that *Cxcl14* has the lowest Ka/Ks ratio in ligand family, the value even lower than that of housekeeping genes *Gapdh* and *Flnb* in GY model. In the receptor family *CXCR4* the receptor gene of CXCL12 has the lowest Ka/Ks ratio, on the contrary, *CXCR2* exhibited the highest Ka/Ks, indicating a coevolution mode between CXC ligand and receptor.

**Supplementary Figure 8.**
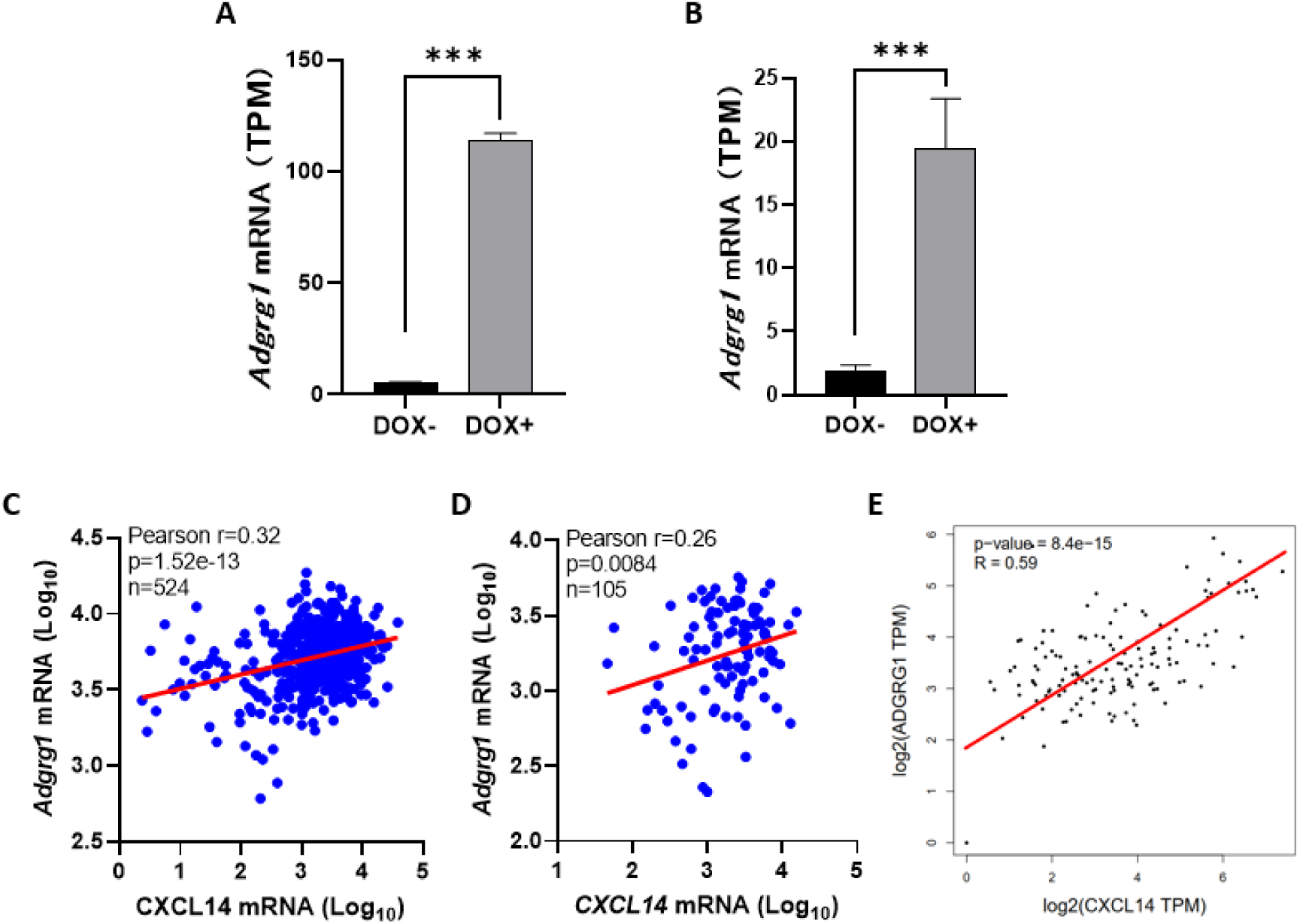
CXCL14 upregulates ADGRG1 mRNA in colorectal carcinoma and normal colon. **(A)** The overexpressed CXCL14 (induced by DOX) significantly enhances the expression of ADGRG1 in MC38 cells grown in vitro. **(B)** The overexpression of CXCL14 (induced by DOX) promotes the expression of ADGRG1 in MC38 cells grown as subcutaneous tumors. **(C)** and **(D)** The database TCGA and CPTAC colon cancer demonstrate a significant positive correlation between CXCL14 and ADGRG1. **(E)** In the GTEx database, a significant positive correlation was observed between CXCL14 and ADGRG1 in normal colon tissue (n=136).

