## Supplemental Table for "CXCL14 suppresses the progression of colon cancer by regulating tumor epithelial-mesenchymal transition and tumor microenvironment"

Supplementary Table 1. Primers for qPCR

| **Primer** | **Sequence (5‘-3’)** | **target** |
| --- | --- | --- |
| mCXCL14-q4s | GCACTGCCTGCACCCTAAG | Endogenous mouse *CXCL14* mRNA |
| mCXCL14-q4a | CTTCGTAGACCCTGCGCTTC |  |
| mCXCL14-Q3s | CAGAGCACCAAACGCTTCATC | Plasmids derived mouse *CXCL14* mRNA |
| mCXCL14-Q3a | TTCTTTCCATGATCGTCCACCC |  |
| hCXCL14-qs | AAGCCAAAGTACCCGCACTG | Endogenous mRNA or plasmids derived human *CXCL14* mRNA |
| hCXCL14-qa | GACCTCGGTACCTGGACACG |  |
| mMmp14-qs | TTCCAATGATCCCTCCGCCA | Endogenous mouse *Mmp14* |
| mMmp14-qa | GACCCTGACTTGCTTCCATAAA |  |
| mFn1-qs | GACGCCGTTCCAGGAGAGTT | Endogenous mouse *Fn1* |
| mFn1-qa | AGTCAGAGTCGCACTGGTAGA |  |
| mTK1-qs | AGCAACAGCTTCTCCACACA | Endogenous mouse *TK1* |
| mTK1-qa | CAAGGACTCCTGGGTCACATC |  |
| mCDH2-qs | TTCTGGCGGCCTTGCTT | Endogenous mouse *CDH2* |
| mCDH2-qa | CGGTAAGACTGCGCTGTAAA |  |
| mCDH1-qs | AACGCTCCTGTCTTCAACCC | Endogenous mouse *CDH1* |
| mCDH1-qa | GGTCACTTTGAGTGTGGCGA |  |
| mTwist1-qs | TTCACAAGAATCAGGGCGTG | Endogenous mouse *Twist1* |
| mTwist1-qa | CTGCCCCTCTGGGAATCTCT |  |
| mVim-qs | AGCACCCTGCAGTCATTCAG | Endogenous mouse *Vim* |
| mVim-qa | TCCACTTTCCGTTCAAGGTCA |  |
| mSnai1-qs | GTCCAGCTGTAACCATGCCT | Endogenous mouse *Snail1* |
| mSnai1-qa | TGTCACCAGGACAAATGGGG |  |
| mSox2-qs | GGAGGAGAGCGCCTGTTTTT | Endogenous mouse *Sox2* |
| mSox2-qa | CTGGCGGAGAATAGTTGGGG |  |
| mOcln-qs | GAACTGTGGATTGGCAGCG | Endogenous mouse *Ocln* |
| mOcln-qa | AGCAAAATGTCCAGGCTCCC |  |
| mMmp9-qs | GAGTTCTCTGGTGTGCCCTG | Endogenous mouse *Mmp9* |
| mMmp9-qa | TTGGAAACTCACACGCCAGA |  |
| mMmp2-qs | ACGATGATGACCGGAAGTGG | Endogenous mouse *Mmp2* |
| mMmp2-qa | GTCCTGAGAGTGTTCCAGCC |  |
| mKrt10-qs | ACGAGAAGCATGGCAACTCA | Endogenous mouse *Krt10* |
| mKrtq0-qa | TTGTCAGGGTGAGGATCTGC |  |
| mFas-qs | CCATGCACAGAAGGGAAGGA | Endogenous mouse *Fas* |
| mFas-qa | GGGTGCAGTTTGTTTCCACC |  |
